# Comparison of Machine Learning Methods in Mild Cognitive Impairment Prediction for Cancer Patients Using EHR Data

**DOI:** 10.1101/2023.02.24.23286413

**Authors:** Xiaodan Zhang, Martin Witteveen-Lane, Yanzeng Li, Omkar Kulkarni, Dave Chesla, Bin Chen

**Author notes:** To whom correspondence should be addressed: BC; DC; XZ. Co-first authors (Xiaodan Zhang, Martin Witteveen-Lane).

## Abstract

Cancer and dementia are common in aging populations. Mild cognitive impairment (MCI) is a stage between the cognitive changes of normal aging and dementia that can lead to a decline in quality of life. With the substantial improvement of survival in many cancers, maintaining a high quality of life has become a new goal in cancer care. Identifying those patients with a high risk of developing MCI may facilitate early intervention and further improve patient care. The objective of this study is to survey machine learning techniques and AutoML to model the early detection of MCI in patients with cancers using the features which are known risk factors in dementia and accessible in the electronic health records (EHR). We compared multiple machine learning methods and explored AutoML to predict 1-year risk of MCI for cancer patients. Among 27 models, XGBoost in AutoML gave the highest AUC (0.79), suggesting the superiority of using automated machine learning tools to search for the best model and parameters. The feature importance analysis revealed that cancer patients with brain malignancy, hypertension, or cardiovascular diseases are more likely to develop MCI. The overall poor performance indicates more efforts should be made to improve data quality and increase features and sample size.

## Introduction

Mild cognitive impairment (MCI) can occur naturally as a normal byproduct of aging, and will sometimes progress towards more serious conditions such as Dementia or Alzheimer’s Disease. MCI or difficulties with memory and attention are one of the top complaints of cancer patients found in the literature^1^. A recent web-based survey revealed 75% of cancer patients reported cognitive complaints related to treatments^2^. As the advances in diagnostic and therapeutic strategies have greatly improved the overall survival in many cancers, seeking high-quality of life including maintaining normal cognitive function is emerging as a tangible goal in cancer care. Predicting susceptibility for MCI, and understanding the progression towards cognitive disease in cancer patients, could be of great benefit to public health as well as contribute to cost-preventative measures in our health care systems.

Cancer and dementia are both common in aging populations, and previous research has suggested inverse^3,4^ or direct^5,6^ associations between the two conditions. The occurrence of cognitive impairment in cancer patients could be due to treatment, age, predisposing genetic factors and comorbidities^7^. The risk factors of MCI are characterized in many studies, although different populations may present distinct risk factors. In recent studies, several machine learning models have been proposed to predict the onset of dementia/MCI^8,9^, however, few studies focused on predicting the onset of MCI for cancer patients, a rapidly growing population, using Electronic Health Records (EHR).

The objective of this study is thus to evaluate different machine learning techniques to model the early detection of MCI in cancer patients using the features collected from EHR data. We leveraged the unique biobank patient data at Corewell Health, the largest health care system in Western Michigan, and explored features that are known risk factors in MCI/dementia. We implemented logistic regression model, random forest model, k-Nearest Neighbor (KNN) model, neural networks (NN) and extreme gradient boosting (XGBoost) model to identify the significant risk factors for MCI, and to predict 1-year MCI risk for cancer patients. The performance of the models was assessed to check if machine learning techniques could be possibly used to predict the MCI risk prior to the onset for cancer patients using EHR data.

In recent years, AutoML has been proposed as a way to expand the application domain of machine learning algorithms and can be deployed in healthcare area ^10^and other variety of fields^10,11^. AutoML provides a framework for domain experts to design machine learning pipelines and carry out model selection and hyperparameter optimization without a deep knowledge of machine learning. Studies have shown that AutoML can accelerate the integration of machine learning in healthcare scenarios^12^, and advance clinical research^13^. H2O.ai is a software platform offering a suite of machine learning and data analysis tools, with AutoML as its automated machine learning solution. We thus explored H2O’s automated algorithm (H2o AutoML) to find the best machine learning model for the 1-year risk of MCI for cancer patients.

## Methods

### Data Source and Collection and Study Design

This study was approved by the Institutional Review Board (IRB) at Corewell Health and Michigan State University (SH 2020-071). The Corewell Health data warehouse stores EHR across 15 hospitals, which contains the digital record of a patient’s health information, such as demographics, diagnoses, medications, and laboratory data. Corewell Health transitioned to Epic from Cerner in 2017, and most of the system shares a single Epic instance. Like many healthcare systems, the combination of legacy data and various hospitals’ coding practices can be challenging. Our team uses a modular scripting approach with SQL to extract data, allowing for easy reuse of code across multiple projects. The Corewell Health’s Biobank is a registry of patient data in Epic, where patients have consented to share their biospecimens and medical records for research purposes. We performed a SQL query on the Corewell Health EHR for subjects, first extracting Biobank patients who were aged 45 and older on the index date. We defined the index date as the MCI diagnosis date for the MCI group, and the last recorded encounter data for the control cohort of cognitively unimpaired (CU) group. The International Classification of Diseases, Ninth Revision^14^ (ICD-9 code = ‘331.83’) and Tenth Revision^15^ (ICD-10 code = ‘G31.84’) were used to identify patients who had a diagnosis of MCI.

Based on the medical reports and the published literature, we then identified 52 major risk factors for MCI, and extracted those risk factors from Epic. When possible, we followed the literature to define and group features. Some features were customized to create a machine-readable dataset (See **Supplementary Table 1** for features definitions). If a feature was not present in a patient’s medical record as indicated by ICD codes, it was assumed to be absent for that patient. Then patients’ EHR records 1 year prior to the index data were pulled out for the prediction. The outcome for the prediction model was defined as the presence or absence of MCI. We excluded Biobank patients without a cancer diagnosis 1 year prior to the index date. Two features with more than 20% missing data were excluded. We excluded patients with missing urbanization (rural, suburban, and urban) group data (0.02% missing values) from analysis. We used mean imputation (MI) for BMI (with 7.56% missing values). Patients’ BMI were categorized as BMI groups (underweight, normal, overweight, and obese), and used in the analysis. Data was gathered up until July 31, 2022. The flowchart of the study is shown in **Figure 1**.

**Figure 1.**
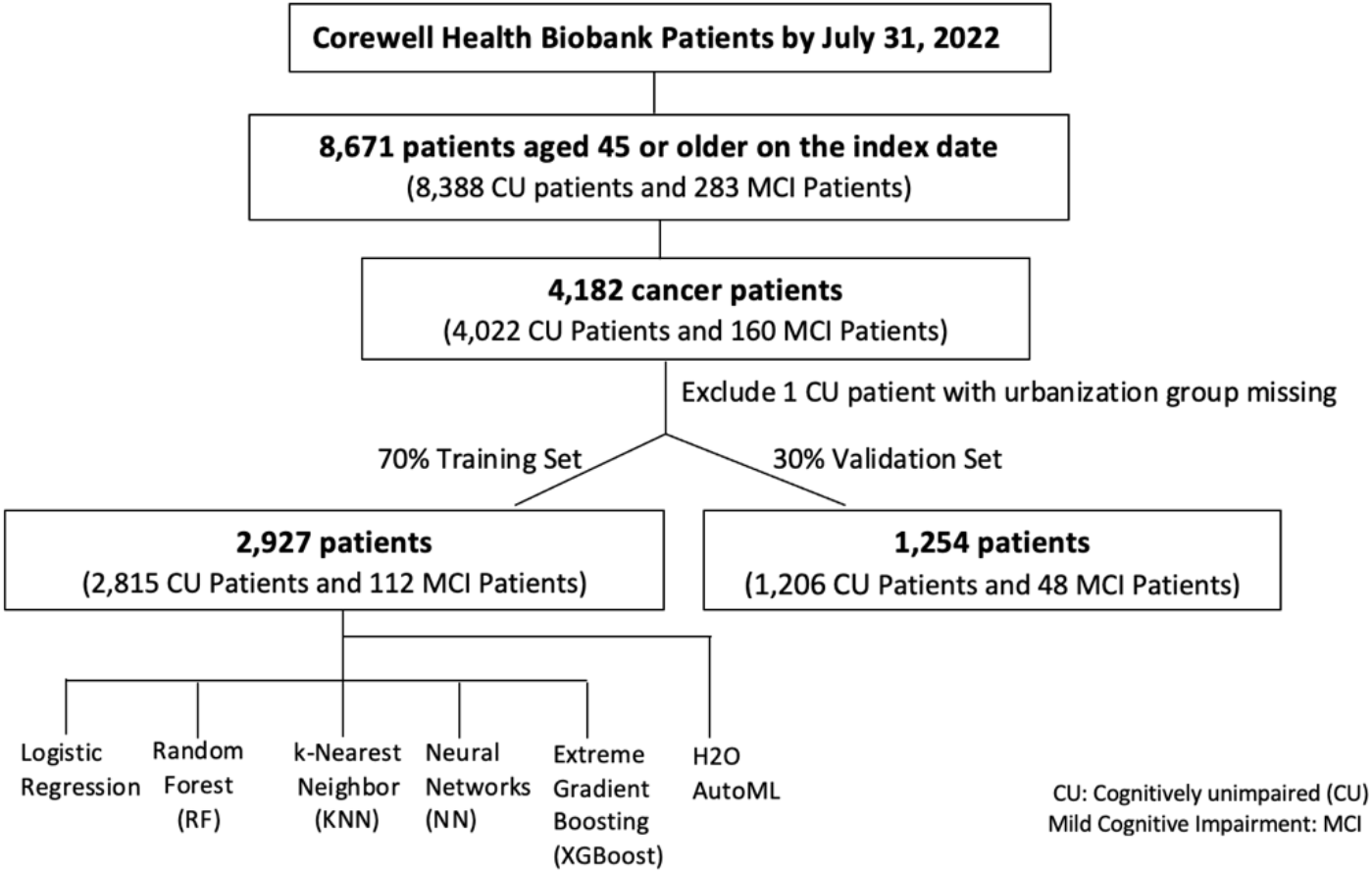
Flowchart of the Machine Learning Modeling for MCI Prediction.

### Statistical Analysis and modeling

The data were split into training (70%), and validation (30%) sets using stratified sampling to train, test and compare different machine learning algorithms. We implemented four baseline machine learning algorithms, including logistic regression, random forest (RF), k-Nearest Neighbor (KNN), Neural networks (NN) and Extreme Gradient Boosting (XGBoost). The algorithms were trained and validated through 10-fold cross-validation. Due to the substantial imbalance between the MCI cases and controls, strategies for resampling, including random under-sampling, random over-sampling, synthetic minority over-sampling technique (SMOTE^16^16) were examined. The SMOTE resampling data well balanced the train set, and has shown the best performance for each machine learning algorithm. However, due to the relatively low sample size, three predictors: acute Leukaemia cancer, TBI, and nasopharynx cancer were removed in the SMOTE resampling, and were not included in the model development. We evaluated the performance of all models by comparing the overall accuracy, sensitivity, specificity, F1 score ((2*((precision*recall)/ (precision + recall))), and area under the receiver operating characteristic curve (AUC). Shapley additive explanation (SHAP)^17^ was used to interpret the results of the best performed model. All the modeling and comparison were performed in R. The caret R package was used to tune the machine learning algorithms. The hyperparameter tunes can be found in **Supplementary Table 3**. R packages including dplyr, ggplot2, tidyr, caret, sjPlot, tableone, InformationValue, DataExplorer, ConfusionTableR and h2o were used.

AutoML was used to create, evaluate and deploy machine learning models using multiple algorithms. The following models were trained and cross validated in H2O AutoML process: three pre-specified XGBoost GBM (Gradient Boosting Machine) models, a fixed grid of GLMs, a default Random Forest (DRF), five pre-specified H2O GBMs, a near-default Deep Neural Net, an Extremely Randomized Forest (XRT), a random grid of XGBoost GBMs, a random grid of H2O GBMs, a random grid of Deep Neural Nets. In addition, a stacked ensemble of all the models trained above, and a “Best of Family” Stacked Ensemble that contains the best performing model for each algorithm class were also developed^18,17^.

## Results

In this study, 8,671 patients aged 45 and older were selected from the Corewell Health Biobank. Out of these, 283 (3%) patients were diagnosed with MCI by July 31, 2022, while 4,182 (48%) patients had a cancer diagnosis in the year prior to the index date. Among the non-cancer group, 123 (2.7%) patients had a diagnosis of MCI. In the cancer group, 160 (3.8%) patients were diagnosed with MCI.

A total of 4,181 patients were included in the final analysis, with a female-to-male ratio of approximately 4:3 (2349 women and 1832 men). Of these, approximately 10.78% (451 patients) were aged 45-54, 25.66% (1,073 patients) aged between 55-64, 35.49% (1483 patients) between 65-74, 22.09% (924 patients) between 75-84, and 5.98% (250 patients) aged 85 and above.

The characteristics of the control group compared with the MCI were shown in **Supplementary Table 2**. Statistically significant differences (*P* value < 0.05) were observed in age group, diabetes, hearing loss, heart disease, hyperlipidemia, hypertension, vascular diseases, antidepressants, CAD, stroke, brain cancer, pancreas cancer, and uterus cancer.

After evaluating the performance of the models, all models showed relatively high specificity but low sensitivity, leading to overall poor F1 scores. The XGBoost model produced the highest AUC of 0.723, with the highest F1 Score (0.170). This model also had the best performance with an accuracy of 0.823, followed by the logistic model with an accuracy of 0.816. The KNN and RF models have an accuracy of 0.794 and 0.784, respectively. The NN model showed the lowest accuracy of 0.767. Overall, the XGBoost model showed a better performance on the accuracy, specificity, F1 score, and AUC compared with other models, however, with a relatively low sensitivity.

### Features importance and explanation of risk factors

Through the importance analysis of variables, the top five most important features for each model were displayed. Antidepressants, vascular disease and brain cancer were the common top features among all the five models. Pancreas cancer, hypertension, hyperlipidemia, and anxiolytics have also been observed as important risk factors in developing MCI.

**Figure 2**a depicts the top 10 features most effective at predicting MCI outcomes in the XGBoost model. The importance matrix plot ranked the variables contributing to 1-year MCI risk prediction from the most to least important as vascular, antidepressants, brain cancer, hyperlipidemia, anxiolytics, age group, BMI group, urbanization, uterus cancer, and colon cancer. In the SHAP summary plot, vascular disease was found to be the most important risk factor, with a high value (presence) in vascular disease corresponding to positive SHAP values and an increase in MCI risk.

**Figure 2.**
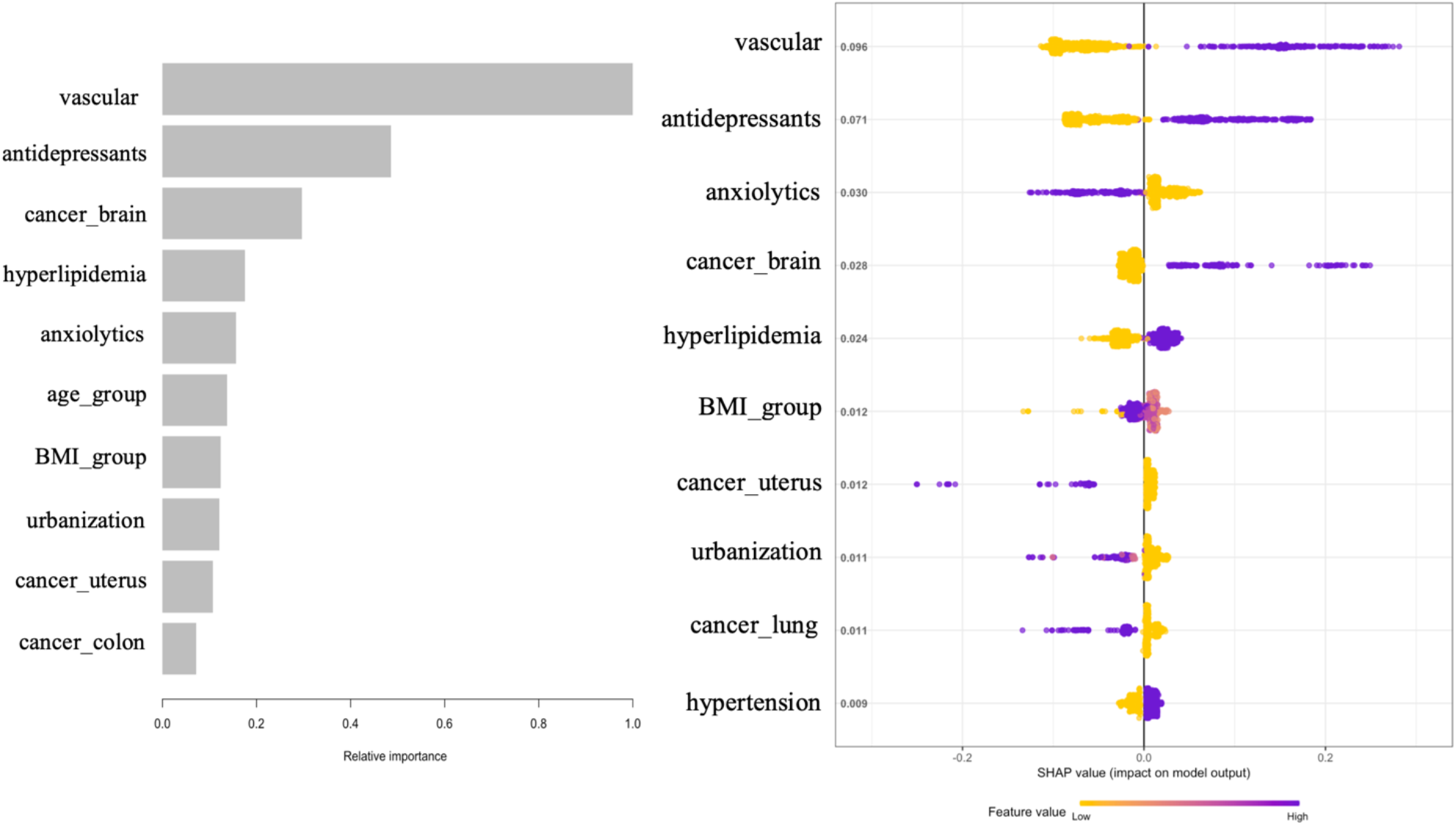
Importance Plot and SHAP Summary Plot for XGBoost model. (a) Importance Plot.(b) SHAP Summary Plot. One dot per patient per feature is colored according to a feature value, where purple represents a higher value and yellow represents a lower value. A positive SHAP value indicates an increase in risk and vice versa.

Similarly, patients with high values (presence) of antidepressants, brain cancer, hyperlipidemia, and hypertension tend to be associated with an increased 1-year risk of MCI. For the features of anxiolytics, uterus cancer, urbanization, and lung cancer, we observed the high values (presence) of those features have negative SHAP values, which indicates that patients with those features were associated with a decreased risk of MCI.

We observed antidepressants, hyperlipidemia, hypertension, and vascular disease shown significant association with increased MCI risk, while patients with anxiolytics use showed a decreased risk. Our findings are consistent with many research outcomes with respect to these significant risk factors. Several studies suggested that antidepressants use was associated with an increased risk of developing MCI or dementia^19,20,21,22^. Hyperlipidemia is an early risk factor for Alzheimer’s disease. Zambón et al found that patients with familial hyperlipidemia showed a high incidence of MCI compared with those without^23^. Previous studies indicated that vascular disease^24,30^ and hypertension^25^ were related to a higher risk of cognitive decline and dementia, including Alzheimer disease (AD). A study conducted by Burke Slet al suggested that anxiolytics might moderate the effect of anxiety on MCI and AD development^26^. Finally, females were at slightly high risk for MCI, corroborating with the observation in Alzheimer’s disease^27^. Although most of the included features were explored to characterize the risk of MCI, our models indicate only a few of them are significant in the cancer population.

### H2O AutoML Exploration

Using H2O AutoML, models were trained, tuned, and cross-validated through the automatic process, with a maximum of 30 models being developed or a maximum running time of 3,000 seconds. A total of 27 machine learning models were implemented to predict the 1-year risk of MCI for cancer patients. The performance of the models was evaluated using AUC values, and the results were displayed in the model leaderboard (**Table 3**). As expected, there is a huge variation of the model performances. Even for the same model, different parameter settings could result in distinct performances, highlighting the necessity to use automated machine learning tools to build baseline models. The best-performing model was identified and shown in the leaderboard with model ID *XGBoost_grid_1_AutoML_2_20230119_91531_model_3*, with the highest AUC of 0.787. This model was selected as the top-ranking model from the list of trained models in the leaderboard. For convenience, we renamed this best model “XGBoost_AutoML”.

**Table 1.** Evaluation of the performance of the five algorithms using SMOTE resampling.

| Model | Accuracy | Sensitivity | Specificity | F1 | AUC |
| --- | --- | --- | --- | --- | --- |
| Logistic | 0.816 | 0.478 | 0.831 | 0.161 | 0.711 |
| RF | 0.784 | 0.522 | 0.795 | 0.155 | 0.703 |
| KNN | 0.794 | 0.458 | 0.807 | 0.145 | 0.710 |
| NN | 0.767 | 0.563 | 0.775 | 0.156 | 0.725 |
| XGBoost | 0.823 | 0.478 | 0.837 | 0.170 | 0.723 |

RF, Random Forest; KNN, k-Nearest Neighbor; NN, Neural Networks; XGBoost, Extreme Gradient Boosting; AUC, Area Under the Receiver-Operating Characteristic Curve.

**Table 2.**
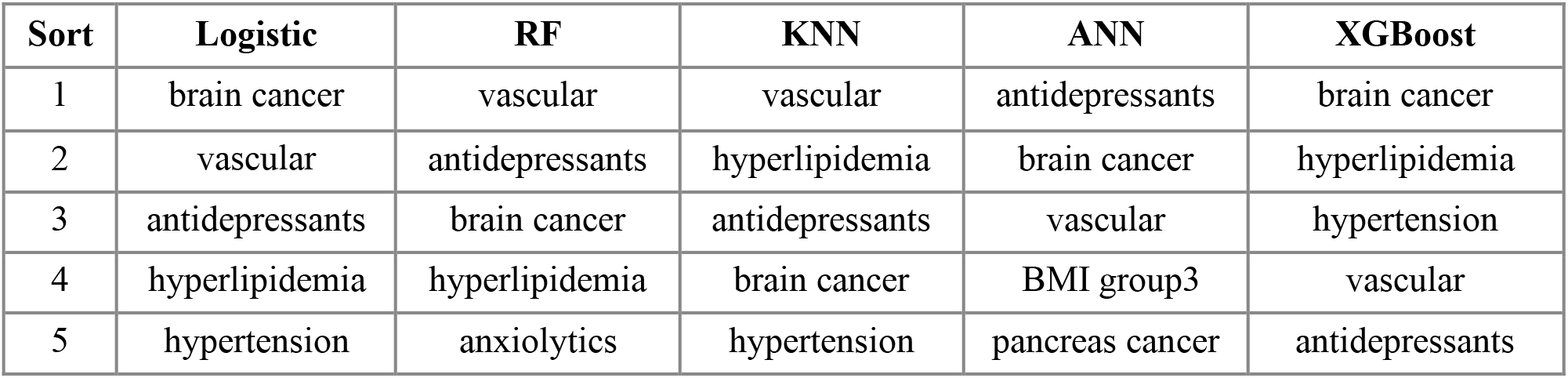
The top 5 significant predictors of the importance of the model.

**Table 3.** Performance Metrics of the Twenty-seven Models in H2O AutoML.

| Rank | Model ID | AUC | AUCPR | logloss |
| --- | --- | --- | --- | --- |
| 1 | XGBoost_grid_1_AutoML_2_20230119_91531_model_3 | 0.787 | 0.147 | 0.142 |
| 2 | XGBoost_grid_1_AutoML_2_20230119_91531_model_1 | 0.786 | 0.139 | 0.143 |
| 3 | XGBoost_grid_1_AutoML_2_20230119_91531_model_4 | 0.785 | 0.134 | 0.142 |
| 4 | GBM_1_AutoML_2_20230119_91531 | 0.783 | 0.144 | 0.146 |
| 5 | StackedEnsemble_AllModels_1_AutoML_2_20230119_91531 | 0.778 | 0.133 | 0.143 |
| 6 | StackedEnsemble_BestOfFamily_1_AutoML_2_20230119_91531 | 0.776 | 0.134 | 0.143 |
| 7 | GLM_1_AutoML_2_20230119_91531 | 0.765 | 0.120 | 0.146 |
| 8 | GBM_grid_1_AutoML_2_20230119_91531_model_4 | 0.764 | 0.110 | 0.152 |
| 9 | XRT_1_AutoML_2_20230119_91531 | 0.756 | 0.101 | 0.159 |
| 10 | XGBoost_grid_1_AutoML_2_20230119_91531_model_2 | 0.751 | 0.124 | 0.152 |
| 11 | GBM_grid_1_AutoML_2_20230119_91531_model_3 | 0.732 | 0.135 | 0.154 |
| 12 | XGBoost_1_AutoML_2_20230119_91531 | 0.730 | 0.095 | 0.264 |
| 13 | DeepLearning_1_AutoML_2_20230119_91531 | 0.728 | 0.100 | 0.175 |
| 14 | DRF_1_AutoML_2_20230119_91531 | 0.723 | 0.089 | 0.203 |
| 15 | DeepLearning_grid_1_AutoML_2_20230119_91531_model_1 | 0.722 | 0.111 | 0.182 |
| 16 | GBM_grid_1_AutoML_2_20230119_91531_model_2 | 0.722 | 0.106 | 0.154 |
| 17 | XGBoost_2_AutoML_2_20230119_91531 | 0.716 | 0.086 | 0.241 |
| 18 | DeepLearning_grid_3_AutoML_2_20230119_91531_model_1 | 0.714 | 0.091 | 0.213 |
| 19 | XGBoost_3_AutoML_2_20230119_91531 | 0.707 | 0.086 | 0.240 |
| 20 | XGBoost_grid_1_AutoML_2_20230119_91531_model_5 | 0.704 | 0.088 | 0.186 |
| 21 | DeepLearning_grid_2_AutoML_2_20230119_91531_model_1 | 0.703 | 0.087 | 0.210 |
| 22 | GBM_grid_1_AutoML_2_20230119_91531_model_1 | 0.701 | 0.087 | 0.162 |
| 23 | GBM_5_AutoML_2_20230119_91531 | 0.700 | 0.087 | 0.161 |
| 24 | GBM_2_AutoML_2_20230119_91531 | 0.696 | 0.081 | 0.166 |
| 25 | DeepLearning_grid_1_AutoML_2_20230119_91531_model_2 | 0.694 | 0.087 | 0.184 |
| 26 | GBM_4_AutoML_2_20230119_91531 | 0.681 | 0.079 | 0.179 |
| 27 | GBM_3_AutoML_2_20230119_91531 | 0.675 | 0.077 | 0.173 |

**Table 4.** Confusion Matrix of XGBoost_AutoML.

|  | CU | MCI |
| --- | --- | --- |
| CU | 1094 | 112 |
| MCI | 28 | 20 |
| Total | 1122 | 132 |

The confusion matrix showed that the accuracy, specificity and F1 score for this best-performing XGBoost_AutoML model were 0.888, 0.975 and 0.222, respectively. The results have been improved comparable with the baseline XGBoost model (0.823, 0.837, 0.170, respectively). However, sensitivity became lower (XGBoost_AutoML: 0.152, XGBoost baseline model: 0.478), likely because additional SMOTE resampling was employed to balance the performance in the baseline model.

## Discussion

In this study, we explored the potential to employ machine learning algorithms and AutoML for 1-year MCI prediction for cancer patients using routine EHR data. As far as we are aware, this is the first to utilize machine learning and AutoML in the MCI prediction for cancer patients. Our analysis revealed that the 1-year risk of MCI for patients with a brain cancer history was high. Patients with lung cancer or uterus cancer tend to have a low risk of developing MCI. For cancer types, like lung cancer, it may be due to those cancer patients not living long enough to develop MCI/dementia, or that the treatment they received altered the trajectory of MCI development. However, more research needs to be done to explore the association between cancer and MCI. The use of anxiolytics may decrease the risk for developing MCI among cancer patients.

H2o AutoML was also utilized to identify the optimal machine learning model, with the XGBoost model being the best. This result was consistent with our findings with improvement on the model accuracy. However, due to a highly imbalanced dataset, the AutoML models were not effective in classifying MCI patients since only the down-sampling and oversampling in H2o AutoML were used. Our research suggests that the optimal approach would involve first searching for the best model and parameters using AutoML, then manually optimizing them (for example, using SMOTE resampling to address imbalanced data).

However, none of the models led to considerable performances in classifying true positive MCI patients. This is likely because only a relatively small number of patients were available for the analysis with the high imbalanced distribution, and only known risk factors were included. The presence of more comorbidities in the cancer patients could be another reason. For example, vascular disease is a common comorbidity for cancer patients^28^, and a risk factor of MCI^29^. That explained the high odds ratio for vascular in the odds ratio table and forest plot (**Supplementary Table 4 and Figure 1)**. To further increase the model performance, more advanced techniques like Grid Search Cross-Validation for hyperparameter optimization and resampling methods such as SMOTE+ENN and SMOTETomek can be explored. However, our preliminary exploration of H2o and a couple of other AutoML tools (data not shown) suggested only improving machine learning algorithms would not boost the overall performance and the future work should be directed towards improving the quantity and quality of the data. Our recent studies demonstrated a data-driven approach could be used to standardize lab measurements^30^ and the state-of-the-art nature language processing tool like BERT has a huge potential to extract useful features from clinical notes^31^. Nevertheless, more efforts should be made to apply machine learning to construct new features to improve the performance of MCI predictions and identify novel risk factors.

This study has some limitations that future research should address. The models were only trained using data from one health care institution and were note validated with external datasets. A larger cohort of cancer patients should be included in future studies. Additionally, this study did not take into account potential time-varying effects. For example, a diagnosis of hypertension was weighted the same whether it was given 1 year or 3 years before an MCI event. Furthermore, more complex EHR features such as cancer stages, metastasis status, lab measurements, imaging data, and medications were not considered in the analysis. To enhance the prediction performance, these limitations should be considered.

### Conclusion

Early detection of cancer patients at risk of developing MCI can facilitate early intervention and further improve patient care. In this study, we explored the potential of using machine learning techniques and AutoML on EHR data to identify the risk factors of MCI for cancer patients, and to build models that could be possibly used in clinical settings to help predict risk of developing MCI. This study provides a useful baseline for future work, and further efforts are necessary to uncover novel risk factors and improve the model’s performance.

## Data Availability

All data produced in the present work are contained in the manuscript

## Contributor Information

XZ and MWL contributed to the conception and design of the study. XZ contributed to the analysis and interpretation. MWL pulled out the EHR data and took responsibility for the integrity and accuracy of the data. YL explored the Neural Networks model of the data. OK participated in the MCI literature search and the early stage of the discussions. DC and BC supervised and managed the project. MWL and BC aided in result analysis and manuscript development. All authors read and approved the final version of the manuscript to be submitted.

## References

1. Baxter, Mary F., Andrea N. Dulworth, and Theresa M. Smith. “Identification of mild cognitive impairments in cancer survivors.” Occupational therapy in health care 25.1 (2011): 26–37.

2. Lange M, Licaj I, Clarisse B, et al. Cognitive complaints in cancer survivors and expectations for support: Results from a web-based survey. Cancer Med. 2019 May;8(5):2654–2663. doi: 10.1002/cam4.2069. Epub 2019 Mar 18. PMID: 30884207; PMCID: PMC6536919.

3. Driver JA, Beiser A, Au R, et al. Inverse association between cancer and Alzheimer’s disease: results from the framingham heart study. BMJ. 2012; 344:e1442.

4. Musicco M, Adorni F, Di Santo S, et al. Inverse occurrence of cancer and Alzheimer disease: a population-based incidence study. Neurology. 2013; 81:322–328.

5. Realmuto S, Cinturino A, Arnao V, et al. Tumor diagnosis preceding Alzheimer’s disease onset: is there a link between cancer and Alzheimer’s disease? J Alzheimers Dis. 2012; 31:177–182.

6. Nudelman KN, Risacher SL, West JD, et al. Alzheimer’s Disease Neuroimaging Initiative: Association of cancer history with Alzheimer’s disease onset and structural brain changes. Front Physiol. 2014; 5:423.

7. Lange M, Joly F, Vardy J, Ahles T, Dubois M, Tron L, Winocur G, De Ruiter MB, Castel H. Cancer-related cognitive impairment: an update on state of the art, detection, and management strategies in cancer survivors. Ann Oncol. 2019 Dec 1;30(12):1925–1940. doi: 10.1093/annonc/mdz410. PMID: 31617564; PMCID: PMC8109411.

8. Bansal, Deepika, et al. “Comparative analysis of various machine learning algorithms for detecting dementia.” Procedia computer science 132 (2018): 1497–1502.

9. Stamate, Daniel, Wajdi Alghamdi, Jeremy Ogg, Richard Hoile, and Fionn Murtagh. “A machine learning framework for predicting dementia and mild cognitive impairment.” In 2018 17th IEEE International Conference on Machine Learning and Applications (ICMLA), pp. 671–678. IEEE, 2018.

10. Feurer, M.; Klein, A.; Eggensperger, K.; Springenberg, J.; Blum, M.; Hutter, F. Efficient and robust automated machine learning. In Proceedings of the Advances in Neural Information Processing Systems, Montreal, QC, Canada, 7–12 December 2015; pp. 2962– 2970.

11. Hutter, F.; Kotthoff, L.; Vanschoren, J. Automated Machine Learning: Methods, Systems, Challenges; Springer: Cham, Switzerland, 2019.

12. Mustafa A, Rahimi Azghadi M. Automated machine learning for healthcare and clinical notes analysis. Computers. 2021; 10(2). https://doi.org/10.3390/computers10020024.

13. Luo G, Stone BL, Johnson MD, Tarczy-Hornoch P, Wilcox AB, Mooney SD, Sheng X, Haug PJ, Nkoy FL. Automating construction of machine learning models with clinical big data: Proposal rationale and methods. JMIR Res Protoc. 2017; 6(8):175. https://doi.org/10.2196/resprot.7757.

14. International Classification of Diseases, Ninth Revision (ICD-9) https://www.cdc.gov/nchs/icd/icd9.htm

15. International Classification of Diseases, Tenth Revision, Clinical Modification (ICD-10-CM) https://www.cdc.gov/nchs/icd/icd-10-cm.htm

16. Chawla NV, Bowyer KW, Hall LO, et al. SMOTE: synthetic minority over-sampling technique. J Artif Int Res. 2002;16(1):321–357.

17. Lundberg, Scott M., and Su-In Lee. “A unified approach to interpreting model predictions.” Advances in neural information processing systems 30 (2017).

18. H2O.ai. H2O AutoML, June 2017. URL http://docs.h2o.ai/h2o/latest-stable/h2o-docs/automl.html.H2Oversion3.30.0.5

19. Chan, Joyce YC, Karen KL Yiu, Timothy CY Kwok, Samuel YS Wong, and Kelvin KF Tsoi. “Depression and antidepressants as potential risk factors in dementia: a systematic review and meta-analysis of 18 longitudinal studies.” Journal of the American Medical Directors Association 20, no. 3 (2019): 279–286.

20. Lee CW, Lin CL, Sung FC, et al. Antidepressant treatment and risk of dementia: a population-based, retrospective case-control study. J Clin Psychiatry. 2016;77:117–22; quiz 122. doi: 10.4088/JCP.14m09580.

21. Goveas JS, Hogan PE, Kotchen JM et al. Depressive symptoms, antidepressant use, and future cognitive health in postmenopausal women: the Women’s Health Initiative Memory Study. Int Psychogeriatr. 2012; 24:1252–1264. doi: 10.1017/S1041610211002778.

22. Wang C, Gao S, Hendrie HC et al. Antidepressant use in the elderly is associated with an increased risk of dementia. Alzheimer Dis Assoc Disord. 2016;30:99–104. doi: 10.1097/WAD.0000000000000103.

23. Zambón D, Quintana M, Mata P, Alonso R, Benavent J, Cruz-Sánchez F, Gich J, Pocoví M, Civeira F, Capurro S, Bachman D, Sambamurti K, Nicholas J, Pappolla MA. Higher incidence of mild cognitive impairment in familial hypercholesterolemia. Am J Med. 2010 Mar;123(3):267–74. doi: 10.1016/j.amjmed.2009.08.015. PMID: 20193836; PMCID: PMC2844655.

24. Lopez OL, Jagust WJ, Dulberg C, et al. Risk Factors for Mild Cognitive Impairment in the Cardiovascular Health Study Cognition Study: Part 2. Arch Neurol. 2003;60(10):1394– 1399. doi:10.1001/archneur.60.10.1394

25. Reitz C, Tang MX, Manly J, et al. Hypertension and the risk of mild cognitive impairment. Arch Neurol. 2007 Dec;64(12):1734–40. doi: 10.1001/archneur.64.12.1734. PMID: 18071036; PMCID: PMC2672564.

26. Burke SL, O’Driscoll J, Alcide A, Li T. Moderating risk of Alzheimer’s disease through the use of anxiolytic agents. Int J Geriatr Psychiatry. 2017 Dec;32(12):1312–1321. doi: 10.1002/gps.4614. Epub 2016 Nov 2. PMID: 27805724; PMCID: PMC5441966.

27. Podcasy JL, Epperson CN. Considering sex and gender in Alzheimer disease and other dementias. Dialogues Clin Neurosci. 2016 Dec;18(4):437–446. doi: 10.31887/DCNS.2016.18.4/cepperson. PMID: 28179815; PMCID: PMC5286729.

28. Wedding U, Roehrig B, Klippstein A, Steiner P, Schaeffer T, Pientka L, Höffken K. Comorbidity in patients with cancer: prevalence and severity measured by cumulative illness rating scale. Crit Rev Oncol Hematol. 2007 Mar;61(3):269–76. doi: 10.1016/j.critrevonc.2006.11.001. Epub 2007 Jan 4. PMID: 17207632.

29. Solfrizzi V, Panza F, Colacicco AM, D’Introno A, Capurso C, Torres F, Grigoletto F, Maggi S, Del Parigi A, Reiman EM, Caselli RJ, Scafato E, Farchi G, Capurso A; Italian Longitudinal Study on Aging Working Group. Vascular risk factors, incidence of MCI, and rates of progression to dementia. Neurology. 2004 Nov 23;63(10):1882–91. doi: 10.1212/01.wnl.0000144281.38555.e3. PMID: 15557506.

30. Liu K, Witteveen-Lane M, Glicksberg BS, Kulkarni O, Shankar R, Chekalin E, Paithankar S, Yang J, Chesla D, Chen B. BGLM: big data-guided LOINC mapping with multi-language support. JAMIA Open. 2022 Nov 25;5(4):ooac099. doi: 10.1093/jamiaopen/ooac099. PMID: 36448022; PMCID: PMC9696745.

31. Liu K, Kulkarni O, Witteveen-Lane M, Chen B, Chesla D. MetBERT: a generalizable and pre-trained deep learning model for the prediction of metastatic cancer from clinical notes. AMIA Annu Symp Proc. 2022 May 23;2022:331–338. PMID: 35854741; PMCID: PMC9285138.

